# Pinna-Imitating Microphone Directionality Improves Sound Localization and Discrimination in Bilateral Cochlear Implant Users

**DOI:** 10.1101/2020.03.05.20023937

**Authors:** Tim Fischer, Christoph Schmid, Martin Kompis, Georgios Mantokoudis, Marco Caversaccio, Wilhelm Wimmer

**Author notes:** Corresponding author: Marco Caversaccio, Department of ENT, Head and Neck Surgery, Inselspital - Bern University Hospital, Freiburgstrasse 4, Bern 3010, Switzerland.

## Abstract

**Objectives:** To compare the sound-source localization, discrimination and tracking performance of bilateral cochlear implant users with omnidirectional (OMNI) and pinna-imitating (PI) microphone directionality modes.

**Design:** Twelve experienced bilateral cochlear implant users participated in the study. Their audio processors were fitted with two different programs featuring either the OMNI or PI mode. Each subject performed static and dynamic sound field spatial hearing tests in the horizontal plane. The static tests consisted of an absolute sound localization test and a minimum audible angle (MAA) test, which was measured at 8 azimuth directions. Dynamic sound tracking ability was evaluated by the subject correctly indicating the direction of a moving stimulus along two circular paths around the subject.

**Results:** PI mode led to statistically significant sound localization and discrimination improvements. For static sound localization, the greatest benefit was a reduction in the number of front-back confusions. The front-back confusion rate was reduced from 47% with OMNI mode to 35% with PI mode (*p* = 0.03). The ability to discriminate sound sources at the sides was only possible with PI mode. The MAA value for the sides decreased from a 75.5 to a 37.7-degree angle when PI mode was used (*p* < 0.001). Furthermore, a non-significant trend towards an improvement in the ability to track sound sources was observed for both trajectories tested (*p* = 0.34 and *p* = 0.27).

**Conclusions:** Our results demonstrate that PI mode can lead to improved spatial hearing performance in bilateral cochlear implant users, mainly as a consequence of improved front-back discrimination with PI mode.

## INTRODUCTION

The pinna transforms the spectral characteristics of incoming sound signals dependent on their distance and direction of arrival. The monaural cues resulting from this transformation are particularly important for up-down and front-back localization (Blauert 1997). Cochlear implant (CI) users wearing behind-the-ear audio processors with integrated microphones are not able to utilize pinna cues, which may result in poor localization performance. It has been demonstrated that artificial imitation of the pinna effect in the form of frequency-dependent microphone directionality improves speech intelligibility in noise of CI users when speech and noise are spatially separated (Chung et al. 2004; Kordus et al. 2015; Wimmer et al. 2016; Honeder et al. 2018; Dorman et al. 2018). The pinna-imitating (PI) microphone directionality mode available in the Sonnet audio processor (MED-EL GmbH, Innsbruck, Austria), combines the signals of two matched omnidirectional microphones to form a fixed directionality pattern with an increased sensitivity towards the front for frequencies above 2 kHz. The microphones are located in a front/back configuration at the top of the speech processor with a spatial separation of 1 cm. Beamforming is achieved by utilization of phase differences in the arriving sound signal. If only the signal of the front microphone is used, an omnidirectional (OMNI) microphone characteristic is obtained.

In contrast to speech intelligibility in noise, only limited data for the localization performance of bilateral cochlear implant (BiCI) users with pinna imitation algorithms are available. Dorman et al. (2018) investigated the influence of a pinna imitation algorithm in BiCI users, concluding no positive effects on sound localization performance. The applied test setup was limited to the frontal azimuth only and did not enable the evaluation of front-back confusions (FBCs). Mantokoudis et al. (2011) placed experimental microphones inside the external auditory canal and measured the minimum audible angle (MAA) of BiCI users. Indeed, sound discrimination at the sides was improved, indicating a potential advantage of pinna cues in BiCI users. This finding is consistent with experiments that showed reduced numbers of FBCs in hearing aid users when applying pinna-imitating microphone directionality (Keidser et al. 2006; Van den Bogaert et al. 2011; Mueller et al. 2012; Kuk et al. 2013; Jensen et al. 2013).

Another important aspect of spatial hearing performance is the ability to track moving sound sources, especially when realistic test conditions are desired. The ability of BiCI users to perceive and track a virtual moving stimulus in the frontal azimuth has been investigated by Moua et al. (2019). They reported poor performance in discriminating the movement direction and observed overshooting when judging the actual range of sound movements. The experiments used nonindividualized binaural recordings and were limited to the frontal azimuth.

To summarize, the spatial hearing of BiCI subjects in general and the influence of the PI directionality in particular remain incompletely understood. Therefore, the aim of this single-blinded study was to investigate the spatial hearing performance of BiCI users in the full azimuthal plane. Since the pinna causes considerable level differences above 2 kHz (pinna shadow) and CI users primarily rely on level cues (Loiselle et al. 2016), we hypothesized that a pinna-effect-imitating microphone setting improves the spatial hearing abilities of BiCI users. As a consequence of the higher sensitivity to sounds arriving from the front, the PI directionality mode should introduce larger changes in ILD magnitude as a function of azimuthal source location, in particular for the front/rear directions. To test our hypothesis, we performed a series of sound field experiments using real sound sources and compared the sound localization, discrimination and tracking abilities of BiCI subjects between the OMNI and PI microphone directionality modes.

## MATERIALS AND METHODS

### Study Design and Participants

This prospective study was designed in accordance with the Declaration of Helsinki and was approved by our local institutional review board (Kantonale Ethikkommission-Bern, Switzerland, No. 2018-00901). The study was classified as an observational study according to the Swiss jurisdiction and was deemed to not require registration as a clinical trial. All the participating CI users were postlingually deafened adults even though in four cases the hearing loss was considered to be congenital/progressive. CI01, CI07 and CI12 used hearing aids since early childhood before receiving CIs sequentially. A detailed summary of the participants is provided in Table 1. Only users with monosyllabic word recognition scores in quiet of 70% or better at 60 dB SPL were included in the study. CI users with asymmetries in the number of active electrodes (i.e., more than 1 electrode difference) were not included. This criterion was chosen to increase the homogeneity of the electrode configurations within our study cohort to enable a comparison of results with continuing studies investigating the influence of temporal fine structure preservation on spatial hearing. As a reference, the data of twelve normal-hearing adults were included (Fischer et al. 2019). The participants gave written informed consent before undergoing the study procedure.

**Table 1:** Overview of the study participants. CI = cochlear implant, F = female, M = male, HL = hearing loss.

| Subject | Age, years | Sex | Etiology | Details for Left CI/Right CI |  |
| --- | --- | --- | --- | --- | --- |
|  |  |  |  | Active electrodes | Experience, years |
| CI01 | 20 | M | Congenital/progressive HL | 12/12 | 5/17 |
| CI02 | 67 | M | Otosclerosis | 10/11 | 16/15 |
| CI03 | 48 | F | Progressive HL | 12/12 | 8/9 |
| CI04 | 62 | M | Progressive HL | 12/12 | 6/1 |
| CI05 | 64 | F | Congenital/progressive HL | 11/11 | 12/13 |
| CI06 | 66 | M | Unknown/progressive HL | 12/12 | 19/17 |
| CI07 | 25 | F | Congenital/progressive HL | 12/11 | 17/13 |
| CI08 | 44 | M | Meningitis | 10/9 | 12/6 |
| CI09 | 58 | F | Progressive HL | 10/9 | 3/6 |
| CI10 | 57 | F | Progressive HL | 12/12 | 12/14 |
| CI11 | 62 | F | Unknown | 12/12 | 7/1 |
| CI12 | 22 | F | Congenital/progressive HL | 12/12 | 4/3 |

### Audio Processor Fitting

Audio processor fitting was performed in a single-blinded, counterbalanced fashion. All the subjects had at least 4 weeks of experience with their audio processors worn behind both ears before participating in the study. As default setting, the audio processors were programmed with a fine structure preserving sampling coding strategy (FS4) and activated PI directionality mode, which corresponds to the standard fitting used in our clinical routine. Before the study, the audio processors were all set to the same automatic gain control with a compression ratio of 3:1. In addition, the wind noise reduction feature was disabled, and the sensitivity settings were set to the same value for both processors. There was no matching of single electrodes in pitch or volume beyond the clinical routine fitting. According to our counterbalanced study design, half of the subjects started with the microphone directionality set to OMNI, while the other half had an activated PI mode. The subjects were not informed about which settings were activated on their audio processor. Before starting new tests, a trial run was performed for training, the results of which were discarded.

### Measurement Setup

The subjects were seated in the center of a horizontal circular loudspeaker setup with a radius of 1.1 m inside an acoustic chamber with an approximate reverberation time of 200 ms for frequencies between 0.25 and 10 kHz. The number of loudspeakers (Control 1 Pro, JBL, Northridge, USA) and their positions were adjusted according to the test procedures. To enable real-time dynamic positioning of loudspeakers at different azimuths, we used loudspeakers that were mounted on wireless controllable carriages (see Figure 1) (Fischer et al. 2019). A sound-transparent curtain was used to avoid visual cues during testing. Each loudspeaker was equalized and calibrated with a free-field microphone (type 4133 and preamplifier type 2639, Brüel & Kjaer, Nærum, Denmark) in the center of the circular setup and an audio analyzer (UPV, Rohde & Schwarz, Munich, Germany). The study participants wore eye-tracking glasses during all tests (Pupil Labs, Berlin, Germany) to monitor the head and gaze positions and to minimize orientation-based systematic errors (Razavi et al. 2007). The entire test setup is shown in Figure 1. See supplemental digital content 1 for a video of the setup during an audiometric measurement. A more detailed description of the measurement setup can be found in the article by Fischer et al. (2019).

**Figure 1:**
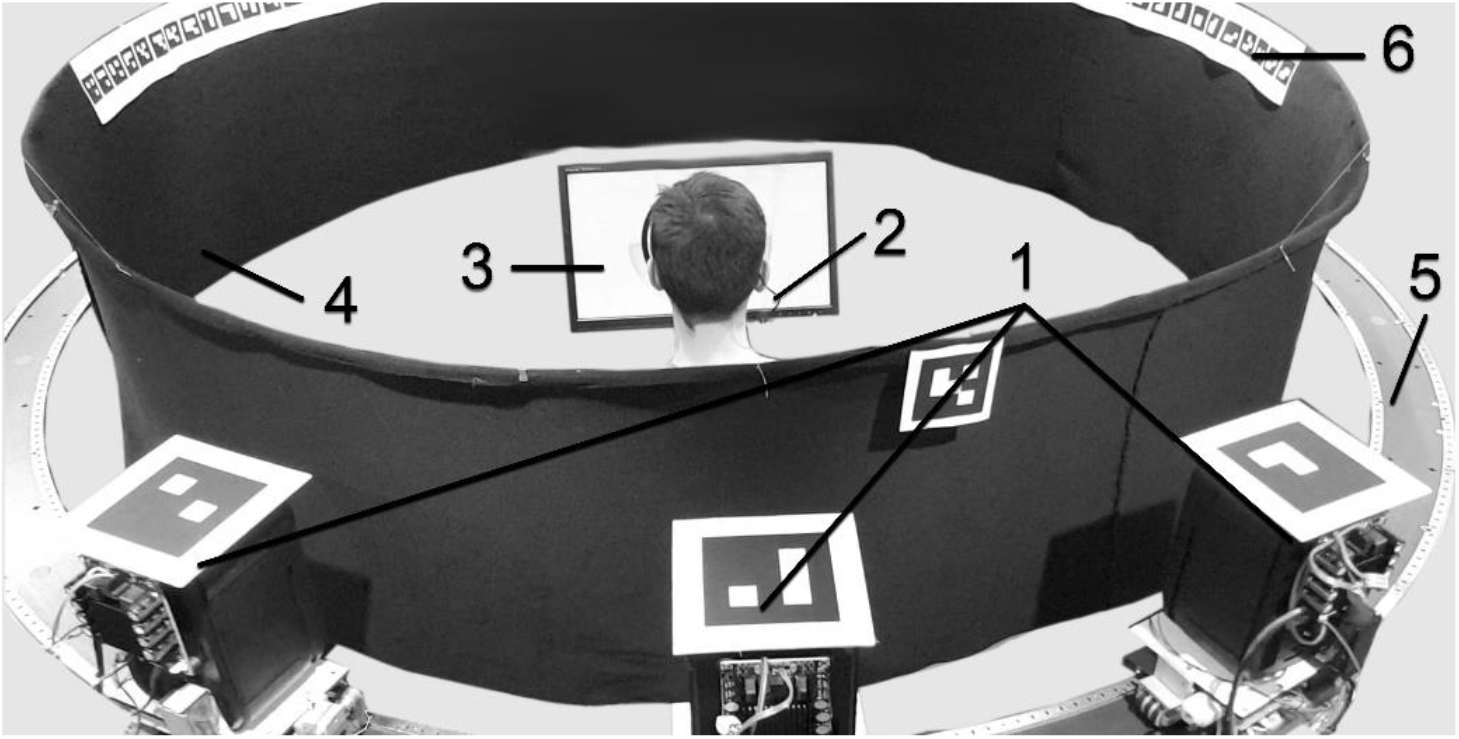
The robotic measurement setup during the minimum audible angle (MAA) assessments at 180-degree azimuth. Three wireless controllable audio robots with optical tracking markers (1), eye-tracking glasses (2), a touch screen with a graphical user interface (GUI) (3), sound-transparent curtain (4), a low-noise rail (5) and azimuth reference markers (6).

### Test Procedures

The following test procedures were performed by the study participants with the OMNI and PI microphone directionality modes: (i) static sound-source localization, (ii) MAA assessment for testing sound discrimination, and (iii) sound-source tracking using graphical user interface (GUI) feedback. To minimize bias effects, a counterbalanced study design was used for the microphone directionality modes, the test procedure order and the individual test items. If desired by the subjects, short breaks were taken at any time. The test procedures are described in more detail in the article by Fischer et al. (2019).

#### Static Sound-Source Localization

We tested the absolute sound localization performance using 12 equally spaced loudspeakers covering the whole azimuthal plane. Three pink noise stimuli with a duration of 200 ms, a 10 ms rise/decay time and a level of 65 dB SPL (roving ±5 dB) were presented from the loudspeakers in a randomized order (Wimmer et al. 2017; Fischer et al. 2019), resulting in a total of 36 stimuli. The subjects had no prior knowledge about the positions of the loudspeakers, and no feedback was given about the correctness of their answers. They were instructed that stimuli could be presented from any azimuth. Subjects indicated the estimated position using a 1-degree-angle resolution dial-on touchpad. Two seconds after a given response, the next stimulus was presented. Before the test, the subjects could familiarize themselves with the test procedure. The test procedures are described in more detail in the article by Fischer et al. (2019).

#### Minimum Audible Angle

To investigate the subjects’ discrimination abilities, the MAA was measured (Mills 1958) at 8 positions, covering the azimuthal plane in 45-degree angle intervals. The MAA is the smallest angular distance that can be detected between the sound sources of two successive tone pulses (Mills 1958). Two tones are played, whereas the second tone is either shifted to the left or to the right regarding the first tone, which stays at the reference position.

For each test step, two pink noise stimuli with a duration of 200 ms (10 ms rise/decay time) at a sound pressure level (SPL) of 65 dB were used, separated by a 1-second intrastimulus interval. The perceived stimulus shift was indicated by the subjects using a touchpad showing a GUI, which adapted to the current measurement position. No feedback was given about the correctness of the answers. Before the test, the subjects could familiarize themselves with the test procedure. The test sequence of the azimuthal test positions (i.e., clockwise vs. counterclockwise) was counterbalanced to minimize bias. The first MAA sequence was tested in the frontal direction (0-degree angle). The test procedures are described in more detail in the article by Fischer et al. (2019).

#### Sound-Source Tracking

For sound-source tracking, a continuous pink noise stimulus with a 65-dB SPL was presented from a loudspeaker that was moved with a wireless robotic setup (Fischer et al. 2019). The subjects were instructed to follow the physically moving stimulus by indicating its position on a touchpad. The same GUI as in the static sound-source localization test was used (see the supplementary material of Fischer et al. (2019)). To familiarize the subjects with the task, a trial run was performed, where the position of the stimulus was visible on the GUI. The trajectory of the trial run consisted of a 450-degree angle-spanning trajectory with a single change in direction at the 45-degree azimuth. The maximum angular velocity of the stimulus was 7.4-degree angle per second, which is regarded as suitable for movement detection (Kourosh & Perrott 1990). The test trajectory consisted of a steady trajectory (900-degree angular span, starting at a 315-degree angle with a single change in direction at the 45-degree angle) and an alternating trajectory (2070-degree angular span, starting at a 0-degree angle with 32 changes in direction each at a multiple of 45-degree angle, where each changing position was approached 2 times clockwise and 2 times counterclockwise). The trial and test trajectories are illustrated with video animations in the supplemental material of Fischer et al. (2019). The test procedures are described in more detail in the article by Fischer et al. (2019).

### Outcome Measures

#### Static Sound-Source Localization

The absolute sound localization accuracy was measured with the root mean square error of the static test (RMSE_LOC_, in degree angle) averaged over the total number of stimuli (N = 36) (Hartmann 1983). For a direction-specific analysis, the RMSE_LOC_ was calculated over the 3 stimuli given from a particular direction. To evaluate the influence of possible FBCs on the performance (Letowski & Letowski 2011), the localization accuracy was also evaluated under the exclusion of responses that crossed the interaural axis with respect to the loudspeaker providing the stimulus. Responses with this pattern are referred to in the literature as FBCs (Rayleigh 1907). The chance levels for the overall localization error (RMSE_LOC_) are 104±8-degree angle (including FBCs) and 79±10-degree angle (excluding FBCs). To further quantify the FBCs, the FBC rate, which is defined as the number of FBCs divided by the number of stimuli presented, was computed. The chance level of the FBC rate for the measurement setup used in this study was 50%±9%.

#### Minimum Audible Angle

We measured the MAA with a two-alternative forced-choice procedure with a 2-down, 1-up rule. Each MAA in a specific direction was measured using 24 steps, determined by an updated maximum likelihood estimation procedure (Shen et al. 2015). The number of steps was determined from pilot tests before the study to ensure the convergence of the MAA. The starting step size of 15-degree angle, a logistic-shaped psychometric function and the parameter range of the angular displacement from 0.5-degree angle to 90-degree angle further characterized the procedure. The MAA for each position was defined as the corresponding 80%-correct threshold of the estimated psychometric function (Senn et al. 2005). The chance level of our MAA test procedure for a specific direction was 83±15-degree angle (24-step sequence with a starting step size of 15-degree angle and maximum likelihood step size adaptation).

#### Sound-Source Tracking

Analogous to the static source localization test, the root mean square error (RMSE) was assessed for every sampled position of the stimulus. We refer to the dynamic position-tracking error as RMSE_*ϑ*_(in degree angle). In addition, the percentage of correctly indicated directions of stimulus movement (clockwise or counterclockwise) was recorded. The error between the indicated and actual stimulus angular velocities was calculated as the RMSE_*ω*_(in degree angle per second). To evaluate whether the subjects lost track of the stimulus, we defined events with high indicated velocities (i.e., greater than 3 times the stimulus velocity) as a relocalization of source (ROS). The occurrence of ROS events (N_ROS_), their duration Δ*t*_*ROS*_ (ms) and the improvement in the tracking error after a ROS event (ΔRMSE_*ϑ*,ROS_ in degree angle) were recorded. A detailed description of the metrics is provided in the article by Fischer et al. (2019).

#### Statistical Analysis

Descriptive statistics were used to compare the performance of the subjects on the group level. The effects of the different microphone directionality modes (PI vs. OMNI) were compared using two-sided Wilcoxon signed-rank tests. On the subject level, a correlation analysis was performed by means of Pearson correlation coefficients to assess the relation between the outcome measures and the years of CI experience, word recognition scores, and age of the subjects. The chance levels for the used test procedures were estimated using Monte Carlo simulations. Statistical calculations were performed in MATLAB (version R2018a, The MathWorks Inc., USA).

## RESULTS

### Static Sound-Source Localization

Figure 2 shows the absolute localization accuracy for each stimulus direction for the OMNI and PI modes (see supplemental digital content 2, Figures 1 and 2 for confusion matrices). Under the exclusion of the FBCs, the localization performance in OMNI mode was the worst in the rear azimuth at 150-, 180- and 210-degree angles with RMSE_LOC_ values of 42±18-degree angle, 41±27-degree angle and 44±27-degree angle, respectively. The best localization performance was observed at a 120-degree angle and a 240-degree angle with 17±9-degree angle and 15±9-degree angle, respectively. In PI mode, the localization errors at the rear azimuths (150, 180 and 210-degree angles) were reduced, leading to a similar performance compared to the frontal azimuths (330, 0 and 30-degree angles). Analogous to the results in OMNI mode, the best localization performance was observed at a 120-degree angle and a 240-degree angle (12±8-degree angle and 13±9-degree angle, respectively).

**Figure 2:**
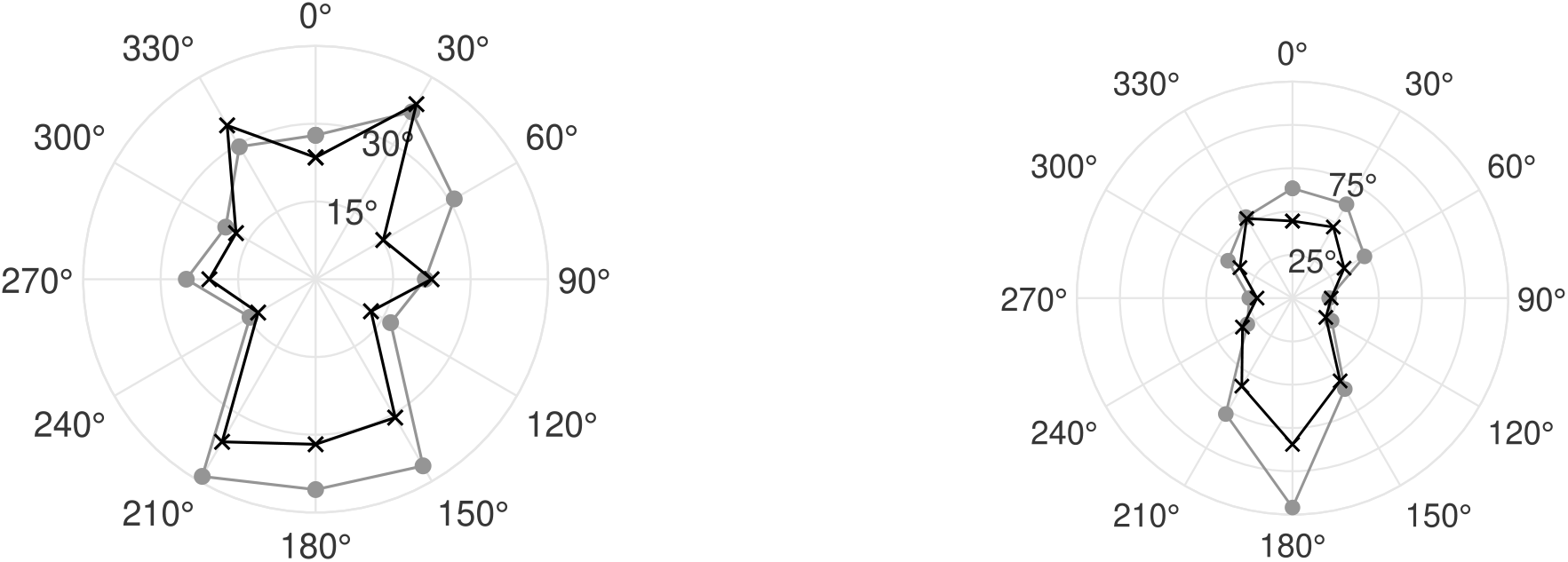
Averaged root mean square error (*RMSE*_*LOC*_) for the omnidirectional (OMNI, circles) and pinna-imitating (PI, crosses) microphone modes. Left, Front-back confusions (FBCs) were excluded from the error calculation whereas the right figure shows the *RMSE*_*LOC*_ including FBCs. See supplemental digital content 2, Tables 2 and 4 for numerical data including standard deviations and median values.

Including the FBCs, for both microphone modes, the lowest RMSE_LOC_ values were observed at a 90-degree and a 270-degree azimuth. Since the FBCs are undefined at these azimuths and do not contribute to the error measure, this finding may not indicate good localization performance, but rather shows the dominance of the FBC related errors. The largest impact of the FBCs on the RMSE_LOC_ was measured at a 180-degree azimuth. See supplemental digital content 2, Tables 1-4 for a detailed summary of the results.

**Table 2:** Summary of outcome measures for the alternating trajectory source tracking experiments. SD = standard deviation, RMSE = root mean square error, ROS = relocalization of source.

|  | Omnidirectional mode |  | Pinna-imitating mode |  |
| --- | --- | --- | --- | --- |
|  | Mean | SD | Mean | SD |
| Position tracking error, $RMSE_{\theta}$ (degree angle) | 67 | 16 | 61 | 20 |
| Velocity tracking error, $RMSE_{\omega}$ (degree angle) | 2.9 | 0.8 | 2.9 | 0.9 |
| Correctly indicated direction (%) | 50 | 5 | 51 | 4 |
| Number of ROS events, $N_{ROS}$ | 25 | 17 | 24 | 15 |
| ROS duration, $\Delta t_{ROS}$ (ms) | 858 | 241 | 817 | 192 |
| Improvement after ROS, $\Delta RMSE_{\theta, ROS}$ (degree angle) | 32 | 10 | 34 | 9 |

The RMSE_LOC_ over all directions with FBCs included was significantly lower in PI mode than in OMNI mode (reduction from 65±7-degree angle to 52±13-degree angle, *p* = 0.007). For comparison, normal-hearing subjects achieved an RMSE_LOC_ of 13±4-degree angle in the identical test procedure (Fischer et al. 2019). Without the FBCs, the improvement in PI mode compared to OMNI mode was not statistically significant (28±6-degree angle to 31±7-degree angle, *p* = 0.29).

The FBC rate improved from 47% (OMNI) to 35% (PI) with *p* = 0.03. As a reference, normal-hearing subjects had an FBC rate of 4%. A separate analysis between front-back and back-front confusions did not show a preference of the PI mode to reduce either front-back or back-front confusions (see supplemental digital content 2, Tables 5 and 6). Of the 12 subjects, 8 had lower FBC rates with the PI mode than with the OMNI mode.

### Minimum Audible Angle

The MAA results for the OMNI and PI modes averaged across all subjects are shown in Figure 3. In both modes, the MAA was best at the front at 0-degree angle and the back at 180-degree angle. The worst performance was measured at the sides at 90 and 270-degree angle. At the sides, where the MAA is equivalent to a front/back discrimination task, a significant improvement from 75.5±23.0-degree angle in OMNI mode to 37.7±21.3 degrees in PI mode (*p* < 0.001) was measured. Furthermore, the MAA averaged across all the measurement directions improved significantly from 30.3±7.6 degrees to 20.8±6.6-degree angle when PI mode was used (*p* = 0.003). For comparison, normal-hearing subjects achieved an averaged MAA of 3.6±1.6-degree angle (Fischer et al. 2019). See supplemental digital content 2, Tables 7 and 8 for individual MAA results with the OMNI and PI modes. Using the OMNI mode, 7 subjects were better than the chance level at the 90-degree angle and the 270-degree angle measurement positions. Two subjects were close to the chance level at the sides. In PI mode, 10 subjects were better than the chance levels. The 2 subjects that did not perform better than the chance level in PI mode, exceeded it for either the 90-degree angle position or for the 270-degree angle position. For both modes, the MAA was significantly smaller for the frontal azimuths (315, 0 and 45-degree angles) than for the rear azimuths (135, 180 and 225-degree angles) (OMNI *p* < 0.001 and PI *p* = 0.01). This could not be observed for the left (225, 270 and 315-degree angles) and right azimuths (45, 90 and 135-degree angles) (OMNI *p* = 0.68 and PI *p* = 0.43). Similar to the absolute localization performance, the correlation analysis did not reveal any statistically significant relations.

**Figure 3:**
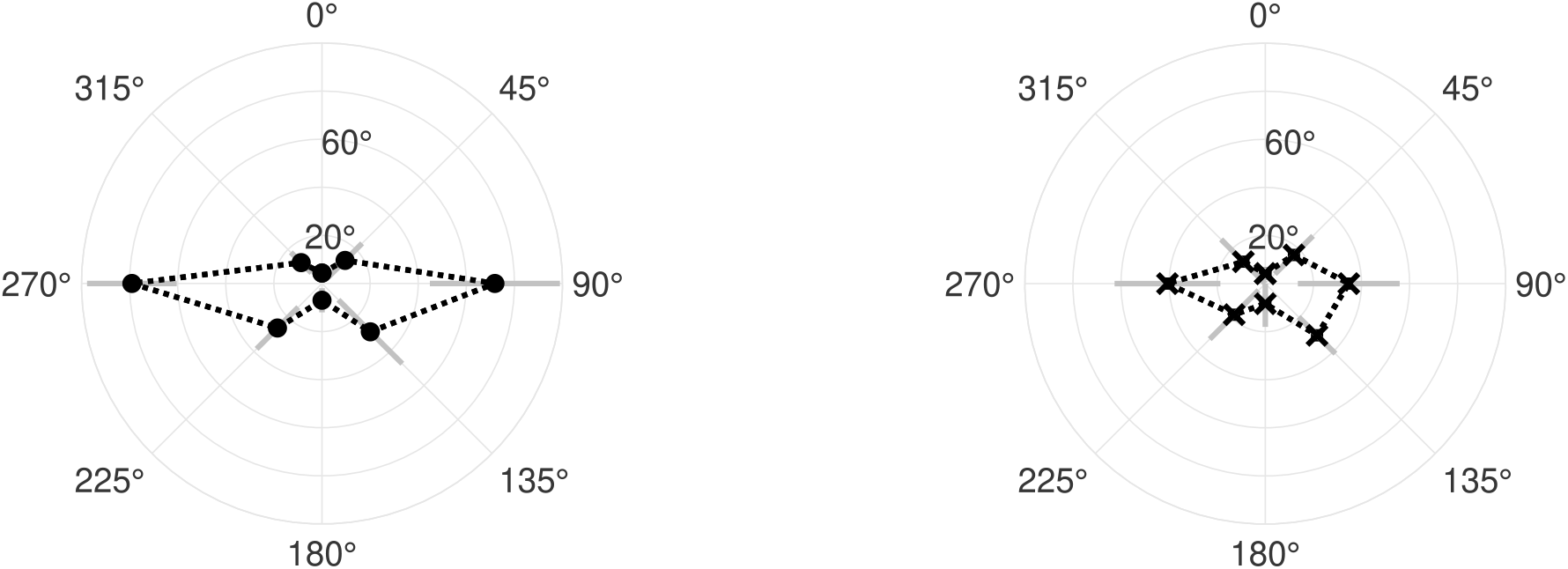
Minimum audible angles for the omnidirectional (OMNI, left figure) and pinna-imitating (PI, right figure) microphone modes averaged across all subjects. The gray bars indicate ±1 standard deviation. See supplemental digital content 2, Tables 7 and 8 for a tabular summary.

### Sound-Source Tracking

#### Steady Trajectory

The subjects had slightly better sound-source tracking abilities with PI mode compared to the performance with OMNI mode. The tracking error RMSE_*ϑ*_and velocity-tracking error RMSE_*ω*_were nonsignificantly reduced from 62.3±12.7-degree angle to 55.6±20.9-degree angle (*p* = 0.34) and from 3.2±1.1-degree angle per second to 2.6 ±1.1-degree angle per second (*p* = 0.20), respectively. The proportion of correctly indicated movement directions throughout the experiments improved nonsignificantly from 64% to 74% (*p* = 0.08). In OMNI mode, 4 subjects did not perceive the single change in direction, and 1 subject indicated a movement in the counterclockwise direction only. This finding was also the case in the PI mode, where 5 subjects did not perceive the change in direction, 2 of them showing an identical tracking behavior as in OMNI mode. It is noticeable that the subjects often used the passages at 0-degree angle or 180-degree angle as the orientation instead of the change in direction (see supplemental digital content 2, Figure 5 for an illustration). For the individual tracking results and outcome measures of the subjects, please see supplemental digital content 2, Tables 9 and 10.

#### Alternating Trajectory

Figure 4 shows the physical sound-source positions, the positions indicated by the subjects and the resulting tracking errors during the alternating-trajectory experiments. The tracking error RMSE_*ϑ*_of the subjects was nonsignificantly reduced from 67-degree angle in PI mode to 61-degree angle (*p* = 0.27) in OMNI mode (see Table 2). As a reference, the normal-hearing subjects achieved an RMSE_*ϑ*_of 19-degree angle in the same experiment (Fischer et al. 2019). On the subject level, 8 subjects achieved a lower RMSE_*ϑ*_with PI mode. For the remaining metrics, the tracking results were similar for both microphone modes. See supplemental digital content 2, Tables 11 and 12 for a detailed summary of the outcome metrics at the subject level.

**Figure 4:**
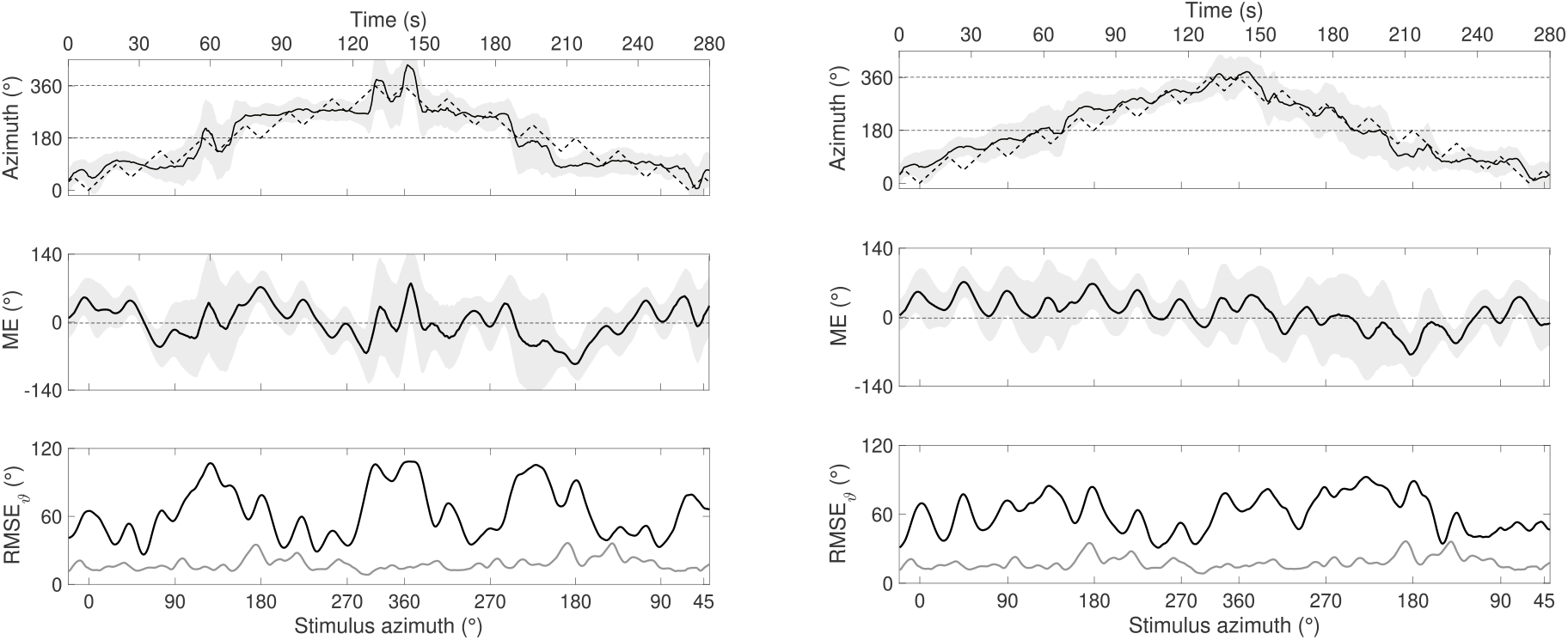
Sound-source tracking with the alternating movement trajectory including multiple changes in direction. Plotted are the positions of the sound source and the averaged subject responses (as indicated with the touchpad), the mean error (ME) and the root mean square error (RMSE_*ϑ*_) averaged across all the subjects for the omnidirectional (OMNI, left figure) and pinna-imitating (PI, right figure) microphone modes. The nonhorizontal dotted lines in the top plots show the trajectory of the stimulus, and the solid lines show the averaged stimulus position, which was indicated by the user. The gray margins around the solid lines in the top and middle plots indicate ±1 standard deviation. The gray lines in the bottom plots indicate the performance of the normal-hearing subjects (Fischer et al. 2019).

Compared to the normal-hearing subjects, BiCI users had great difficulty reacting to changes in direction. Similar to the steady-trajectory experiment, the 0-degree angle and 180-degree angle passages of the sound source were used for orientation. This difficulty was especially visible in OMNI mode, resulting in tracking plateaus when the sound source was in the proximity of the interaural axis (see Figure 4). The movement direction of the sound source (clockwise vs. counterclockwise) did not have an effect on the tracking error RMSE_*ϑ*_.

With both microphone modes, the subjects had a lower position-tracking error (RMSE_*ϑ*_) in the frontal hemisphere than in the rear hemisphere. This difference was statistically significant in PI mode (66±24-degree angle in the back vs. 53±18-degree angle in the front; *p* = 0.02). In OMNI mode, a smaller nonsignificant difference was observed (67±21-degree angle in the back vs. 63±20-degree angle in the front; *p* = 0.60). For comparison, normal-hearing subjects showed a better tracking performance in the front (16±4-degree angle) compared to the back (21±6-degree angle, *p* = 0.005), similar to that in PI mode (Fischer et al. 2019). See supplemental digital content 2, Tables 13 and 14 for additional data of the hemifield and movement direction-related analyses. Only in PI mode did the static localization error RMSE_LOC_ correlate significantly with the corresponding position-tracking error RMSE_*ϑ*_of the alternating trajectory (*p* = 0.03). See supplemental digital content 2, Figure 7 for the scatter plot. Otherwise, the correlation analysis showed no statistically significant correlations in the tracking experiments.

## DISCUSSION

Herein, we present a comprehensive analysis of the sound localization, discrimination and tracking performance of the BiCI users. Our main finding is that the PI microphone directionality mode enables better differentiation between acoustic cues from the front and the back. With PI mode enabled, the BiCI users benefited from the monaural cues provided by the direction-dependent frequency shaping of the pinna-imitating microphone setting (Blauert 1997). As a consequence, the spatial hearing abilities of the participants assessed in terms of sound localization, discrimination and tracking were better when using PI mode than OMNI mode.

A critical examination of the study results or design could raise the question of whether the measured effects were significantly influenced by the different acclimatization periods of the BiCI users between the PI and OMNI settings. However, compared to the experiments performed by Hofman et al. (1998) and Van Wanrooij & Van Opstal (2005), we did not apply a spectral perturbation of the pinna but completely eliminated its influence by applying the OMNI setting, which implied that very little monaural information was available that could have been learned (Opstal 2016). Since our main outcome in this study was to show the ability of PI mode to resolve acoustic ambiguities due to the cone of confusion, we hypothesize that a longer acclimatization period would not significantly alter the measured effect.

### Static Sound-Source Localization

The most obvious influence of PI mode on static sound-source localization is the reduction in the occurrences of FBCs. This reduction resulted in a statistically significant improvement in the overall sound localization error RMSE_LOC_ compared to OMNI mode. Only limited data are available for tests performed with BiCI users in the full horizontal plane, which is required for an analysis of FBCs.

Compared to the study by Pastore et al. (2018), who did not specify a microphone directionality setting and used a setup with coarse angular spacing (60-degree angle) with 3 seconds of stimuli, our observed FBC rates were 7% lower in PI mode and 5% higher in OMNI mode. In contrast to Pastore et al. (2018), the majority of confusions that we observed were not back-to-front confusions. The study by Frohne-Büchner et al. (2004) reported a significant improvement in the localization performance with an in-the-canal microphone for unilateral CI users. However, an FBC analysis was not performed, although they most likely caused the significance in the RMSE_LOC_ differences. Majdak et al. (2011) tested spatial hearing performance with virtual cues presented via the auxiliary inputs of the audio processors and reported an FBC rate of 37%, which was 2% higher than our results with the PI setting. In contrast to this study, the patients received visual feedback after indicating the perceived sound location.

For the frontal azimuth, our results are comparable to the data of Dorman et al. (2018) and Jones et al. (2016), who reported an RMSE_LOC_ of 19-degree angle and 25-degree angle for OMNI mode, respectively. For PI mode, RMSE_LOC_ values of 20-degree angle and 29-degree angle were reported, respectively. In our setting, an RMSE_LOC_ of 27-degree angle for OMNI mode and 23-degree angle for PI mode for the frontal azimuth was observed. On average, no significant differences in the localization performance between the OMNI and PI modes were observed for the frontal hemisphere in our study.

Independent of the microphone setting and including the FBCs in the error calculations, the highest averaged errors for the BiCI users occurred in the very frontal and back azimuths and were rather small at the lateral azimuths. This finding is in line with the available interaural time difference (ITD) and, in particular, ILD cues, which are most prominent at the lateral azimuth. A similar pattern was observed in the study by Majdak et al. (2011). In line with the azimuth-dependent intensities of the binaural cues, the histograms (see supplemental digital content 2, Figures 3 and 4) show that for both microphone settings, the BiCI users tended to perceive the direction more at the sides than at the front or the back. From a statistical point of view, such a response distribution favors the small errors we measured at the sides. In contrast to the findings of Nopp et al. (2004) and Majdak et al. (2011), on average, our study participants had no preference for perceiving stimuli at the left or right azimuth. Additionally, no preference for the frontal versus rear azimuth was observed, regardless of the microphone setting. For this comparison, we excluded the 0-degree angle and the 180-degree angle for the sides’ analysis and the 90-degree angle and the 270-degree angle for the front versus rear analysis. See supplemental digital content 2, Table 16 for a comparison of the data from this study with the references mentioned in this section.

### Minimum Audible Angle

We present an MAA profile of BiCI users for a full azimuth configuration at 8 different directions up to the theoretical measurement limit of ±90-degree angle. Previous studies investigating the MAA of BiCI users were restricted to a maximum displacement between the sound sources of 45-degree angles and thus could not measure the MAA at the sides (Senn et al. 2005; Mantokoudis et al. 2011). To the best of our knowledge, this is the first study to cover MAA values at the sides (i.e., at a 90-degree angle and a 270-degree angle) in BiCI users. With OMNI mode, the majority of our participants had MAAs at the left and right sides that by far exceeded 45-degree angle, which was also observed by Senn et al. (2005) and Mantokoudis et al. (2011). As hypothesized by Senn et al. (2005), the spatial discrimination at the sides was substantially reduced in PI mode. In the study by Mantokoudis et al. (2011), experimental microphones were placed inside the ear canal, demonstrating a theoretical benefit of pinna cues for the sound discrimination of BiCI users at the sides. As our setup was not limited to a maximum MAA of 45-degree angle, we were able to quantitatively show this effect and reproduce the results of Mantokoudis et al. (2011) in a more refined setting.

Since there are no ITD or ILD cues for MAA measurements at the sides, the MAA measurement equals a front/back discrimination task at the 90 and 270 degree angle positions. Similar to the static sound localization tests, the impact of PI mode on the localization or discrimination performance was highest when resolving ambiguities from the cone of confusion. However, we could not observe statistically significant correlations at the subject level between the FBCs in the static sound localization test and the performance of the MAA at the sides with either microphone mode.

We were further unable to confirm the relationship between the MAA at a 0-degree angle and frontal sound-source localization accuracy found by Grieco-Calub & Litovsky (2010) for children using BiCI. In contrast to Senn et al. (2005), we observed statistically significant differences between the frontal quadrants (i.e., at a 45-degree angle and a 315-degree angle, *p* = 0.01) and rear quadrants (i.e., at a 135-degree angle and a 225-degree angle, p <0.001) with both settings. The results for the frontal quadrants were in the range of the results reported by Senn et al. (2005); however, for the rear quadrants, we observed larger MAAs. The difference in performance between the left and right sides was not obvious in our static localization test (i.e., localization errors at 300, 330, 30, 60-degree angles versus at 120, 150, 210, 240-degree angles). For the positions in the front and the back (0 and 180-degree angles, respectively), we could confirm the values of Senn et al. (2005) but report slightly higher values for the back position and OMNI mode due to the high MAA values of 3 subjects. See supplemental digital content 2, Table 15 for a comparison of the data from this study with the references mentioned in this section.

### Sound Source Tracking

The BiCI users had an overall improved sound-source tracking performance with PI mode compared to the OMNI mode; however, the differences in the outcome measures were not statistically significant. The main findings for the alternating- and steady-trajectory experiments are identical regarding the performance of the BiCI users. The continuous movement of the steady trajectory did not provide a reliable sound tracking cue for the BiCI users. Although the tracking error RMSE_*ϑ*_was lower in PI mode than in OMNI mode, the variability between the subjects was higher in PI mode. Compared to our normal-hearing control subjects, the tracking error RMSE_*ϑ*_of the BiCI users was 4 times larger in PI mode (Fischer et al. 2019).

The analysis of the tracking errors using the FBC rate showed a statistically significant lower rate with PI mode (33%±16%) than with OMNI mode (46%±15%, *p* = 0.02). Because the steady trajectory contained no changes in the movement direction of the stimulus at the interaural axis, this effect was only significant for the alternating trajectory. Evaluated in the dynamic setting, FBCs indeed occur more often if pinna-cues were absent. This finding is novel for BiCI users, and we hypothesize that it is a consequence of the high MAA values at the sides. The very frontal (0-degree angle) and backward (180-degree angle) directions seem to be important landmarks for dynamic auditory perception, which can be seen in the “ plateau” tracking in OMNI mode (see Figure 4, left side).

For both of the tested trajectories, the difference in ILD cues had to be evaluated to make an assumption about the shift in the sound source. As the changes in the ILD intensities at the sides are particularly small, we hypothesize that in addition to the cone of confusion, this difference affected the discrimination abilities at the sides. In addition, we observed intense relocalization reactions to the indication of the stimulus position whenever the stimulus passed the sagittal plane, where the difference in ILD cues was at a maximum. On the other hand, we observed a moderate correlation between the RMSE_LOC_, which evaluates absolute ILD cues rather than the differences, and the RMSE_*ϑ*_in the alternating trajectory (see supplemental digital content 2, Figure 7 for the scatter plot). Therefore, not only relative but also absolute localization cues may be of importance for the sound tracking abilities of BiCI users.

Since studies on the continuous tracking ability of BiCI users with regard to a moving stimulus were sparse, a comparison with the literature was only possible to a limited extent. Studies with moving sounds and BiCI users have been performed by Moua et al. (2019) at the frontal azimuth. However, feedback was always provided after the moving stimulus was presented via nonindividualized binaural recordings. Nevertheless, in the study by Moua et al. (2019), as in this study, substantial differences were found in the perception of moving sound sources between BiCI and normal-hearing (NH) subjects. For the alternating trajectories, we observed that BiCI users performed near the chance level of 50% concerning the indication of the correct movement direction. In contrast, normal-hearing subjects achieved a score of 83% (evaluated by Fischer et al. (2019)). For the steady trajectory, a similar pattern was observed with an average performance of 70% correct for the BiCI users and 97% correct for the normal-hearing subjects. The tracking error RMSE_ϑ_ in both trajectories were 3-4 times higher for the BiCI users than for the NH subjects. The continuous presentation of a slowly moving stimulus did not provide a reliable auditory cue for tracking the motion. In addition to sudden changes in the perceived moving direction of the stimulus, it was observed that almost half of the BiCI users, with 2 subjects overlapping in both settings, were not able to detect the only change in direction appearing at a 45-degree angle during the steady-trajectory test. Analyzing the subjects who perceived the change in direction in PI mode and in OMNI mode did not show a significant difference in the perception in terms of reaction times.

As BiCI users almost entirely rely on ILD cues for sound localization in the azimuthal plane (Aronoff et al. (2010)), automatic gain-control settings of the audio processors might play an important role in auditory motion perception. Archer-Boyd & Carlyon (2019) investigated this effect using head movements of a head-and-torso simulator. With the investigated system (Advanced Bionics Ltd., Valencia, CA), they reported that the ILDs received by the BiCI were distorted due to auditory motion induced by head movements. The system used in our study has 4 times smaller compression ratios compared to the devices tested by Archer-Boyd & Carlyon (2019). Nevertheless, automatic gain-control could have caused the limited sound-source tracking performance of the BiCI users. Further investigations with dynamic sound-source positions need to be performed to evaluate this effect.

Because CI processors are not synchronized, different peaks might be picked up by the audio processors, leading to a nonsmooth or erroneous representation of the auditory motion image (Kan et al. 2018). However, as stated above, we believe that automatic gain-control settings, which distort the perception of dynamic ILDs, play a more important role in the perception of auditory motion for BiCI users. We observed that a pinna-imitating microphone setting may help to mitigate distorted ILDs by adding localization cues due to the spectral filtering of incoming sounds.

## Conclusions

In this study, we showed the benefits of pinna-imitating (PI) microphone directionality for the spatial hearing abilities of BiCI users tested under sound field conditions in the horizontal plane. Our results suggest that the PI mode facilitates auditory spatial perception in everyday listening scenarios for BiCI users. Regardless of the microphone setting, BiCI users showed great difficulty in tracking a moving sound source compared to the NH subjects. Furthermore, we hypothesize that the effect of distorted ILD cues due to the automatic gain-control setting could have had an impact on the tracking performance.

## Data Availability

The data referred to in this data is available as supplementary digital content.

## Supplemental Digital Content

- Supplemental Data 1 (Video)
  − SDC_1_Setup_Video.avi
- Supplemental Data 2 (Figures and Tables)
  − SDC2_Figs_and_Tabs.pdf

## Notes

**Conflicts of Interest and Source of Funding** This study was in part funded by the MED-EL Corporation, the Berne University Research Foundation and the Fondation Charidu. There are no conflicts of interest, financial, or otherwise.

### Competing Interest Statement

The authors have declared no competing interest.

### Clinical Trial

KEK-BE, No. 2018-00901

### Funding Statement

This study was in part funded by the Med-El Corporation, the Berne University Research Foundation and the Fondation Charidu.

